# The genetics of hereditary cancer risk syndromes in Brazil: a comprehensive analysis of 1682 patients

**DOI:** 10.1101/2021.04.15.21255554

**Authors:** Jarbas Maciel de Oliveira, Nuria Bengala Zurro, Antonio Victor Campos Coelho, Marcel Pinheiro Caraciolo, Rodrigo Bertollo de Alexandre, Murilo Castro Cervato, Renata Moldenhauer Minillo, George de Vasconcelos Carvalho Neto, Ivana Grivicich, João Bosco de Oliveira Filho

## Abstract

Hereditary cancer risk syndromes are caused by germline variants. Most studies on hereditary cancer have been conducted in white populations. We report the largest study in Brazilian individuals with multiple ethnicities. We genotyped 1682 individuals from all regions of the country with Next-generation sequencing (NGS) panels. Most were women with personal/family history of cancer, mostly breast and ovarian. We identified 321 pathogenic/likely pathogenic (P/LP) variants in 305 people (18.1%) distributed among 32 genes. Most were on *BRCA1* and *BRCA2* (129 patients, 26.2% and 14.3% of all P/LP, respectively), *MUTYH* (42 monoallelic patients, 13.1%), *PALB2* (25, 7.8%), Lynch syndrome genes (17, 5.3%), and *TP53* (17, 5.3%). Transheterozygosity prevalence in our sample was 0.89% (15/1682). *BRCA1/BRCA2* double heterozygosity rate was 0.78% (1/129) for *BRCA* variants carriers and 0.06% (1/1682) overall. We evaluated the performance of the genetic testing criteria by NCCN and Brazilian National Health Agency (ANS). We observed false negative rates of 17.3% and 44.2%, meaning that both failed to detect a substantial part of P/LP positive patients. Our results add knowledge on the Brazilian spectrum of cancer risk germline variants, demonstrate that large multigene panels have high rates of positivity, and indicate that Brazilian testing guidelines should be improved.

## Introduction

Hereditary cancer risk syndromes are a group of disorders caused by germline pathogenic variants in a growing number of genes. They are the main predisposing factor in about 5-10% of all diagnosed cancers. Accurate genetic diagnosis in patients with hereditary cancer risk syndromes may reduce morbidity and mortality by allowing the adoption of specific preventative and therapeutic measures. Additionally, family members at risk can be tested and counseled, extending the clinical benefits to many individuals ^1^.

Currently, there are over 30 hereditary cancer syndromes already described, the most prevalent being hereditary breast and ovarian cancer syndrome (HBOC). Other syndromes include Li-Fraumeni, Cowden, Lynch, Familial Adenomatous Polyposis (APC), multiple Endocrine Neoplasia Type 1 or Wermer’s Syndrome (MEN1), multiple Endocrine Neoplasia Syndrome - Type 2 (RET), and Von Hippel-Lindau Syndrome (VHL), to cite the most common. Most syndromes are inherited in an autosomal-dominant manner, with variable penetrance, and are caused by mutations in oncogenes or tumor suppressor genes ^2^. Although initially hard to diagnose genetically, the advent of next-generation sequencing (NGS) has allowed for testing using panels with dozens to hundreds of genes simultaneously and inexpensively ^3^. This has caused a great increase in the clinical use of these NGS panels, while a growing body of evidence show their clinical utility as compared to single-gene testing ^4, 5^.

Although large studies have been published on the diagnostic utility of such panels, these are mostly restricted to white populations. The aim of this study was to describe the frequency and type of germline pathogenic variants in cancer susceptibility genes in patients with clinical criteria of hereditary cancer syndrome by NGS in the genetically admixed Brazilian population.

## Materials and methods

### Inclusion and exclusion criteria

The cohort to the retrospective study included 1682 Brazilian patients who received a multi-gene NGS panel for risk of hereditary cancer in the Genomika diagnostics unit, a genetics laboratory of the Albert Einstein Israeli Hospital (Genomika-Einstein hereafter). As this study took place in a private diagnostic laboratory this cohort was not specifically selected for any sex, age, ethnicity, or history of cancer. Both patients with or without criteria for hereditary risk cancer syndrome were considered. After genetic assessment we classified patients according to the USA National Comprehensive Cancer Network (NCCN, www.nccn.org) guidelines: genetic/familial high-risk assessment versions 3.2019 and 1.2020. Non-Brazilian patients were excluded.

### Research ethics statement

This project was approved by the Institutional review board from the Research Ethics Committee of the Institute of Integral Medicine Professor Fernando Figueira (CEP-IMIP) in Recife, Pernambuco and all individuals provided written consent for multi-gene testing (protocol number CAAE 29567220.4.1001.0071).

### Multigene genotyping panel

Genotyping was performed through panels that covered the entire coding region including 20 bp flanking each exon and noncanonical splice region. Patients were sequenced with panels that ranged from 27 to 78 genes, including genes mostly associated with hereditary cancer risk, such as *APC, ATM, BARD1, BMPR1A, BRCA1, BRCA2, CDH1, CDK4, CHEK2, EGFR, EPCAM, GREM1 HOXB13, MEN1, MITF, MLH1, MRE11A, MSH2, MSH6, MUTYH, NBN, PALB2, PMS2, POLD1, POLE, PTEN, RAD50, RAD51C, RAD51D, RB1, RET, SMAD4, STK11, TP53, VHL*. The complete list of genes is available in the Supplementary Table 1).

### Library preparation for NGS

Laboratory procedures were performed at Genomika-Einstein units (Recife - PE and São Paulo - SP). Genomic DNA was extracted from peripheral blood or saliva by automation (QIASymphony platform, QIAGEN, Hilden, Germany) using the DNA Mini Kit extraction kit (QIAGEN). The quality and quantity of the extracted DNA were assessed by fluorometry (Qubit, Thermo Fisher, Waltham, Massachusetts). The genomic DNA was enzymatically fragmented and enriched. The fragments were barcoded via multiplexed PCR technology by QIAseq Targeted DNA Panels (CDHS 174272-2274 QIAGEN). The genes were sequenced on MiSeq or Next-Seq 550 instruments (Illumina, San Diego, CA) using MiSeq Reagent Kit v2 (300-cycles) or Next-Seq 550 High-throughput kit (300 cycles) with 100% coverage and 50X minimum depth.

### Sanger sequencing

In cases where variant confirmation was deemed necessary (low read depth, complex indels), the genotype was determined by conventional PCR followed by Sanger sequencing on an ABI 3500 (Thermo Fisher, Waltham, Massachusetts) automatic sequencer. The variant call was made via the reference transcript from the ATG initiation codon aligned against the GRCh37 / hg19 reference genome.

### *Alu* insertion identification

Relevant genetic screenings of breast/ovarian cancer families of Portuguese ancestry demonstrate that the c.156_157ins*Alu* variant is the most frequent *BRCA2* rearrangement. Detection of this variant was performed as published elsewhere ^6^. Briefly, PCR of *BRCA2* exon 3 was performed and the *Alu* insertion is detected by differential agarose gel electrophoresis.

### Variant classification and bioinformatic analysis

Bioinformatic analysis was performed using GATK 3.0 best practices. VCFs were annotated using Annovar and in-house databases. The clinical significance of the variants was determined according to the criteria of the American College of Genomics and Genetics (ACMG) 2015 guidelines for sequence variant interpretation and 2018 guideline update ^7, 8^. Each variant classification were performed independently by two specialists. Allelic frequency was examined through access to population databases as Genome Aggregation Database (gnomAD) ^9^, Exome Aggregation Consortium (ExAC) ^10^, and The 1000 Genomes Project ^11^. Disease information were upload form ClinVar ^12^ and the Leinden Open Variation Database (LOVD) ^13^. All variant information used for classification were updated and upgraded in an in-house local knowledgebase.

## Results

### Study sample characteristics and cancer histories

Between July 2016 and July 2019, we collected samples and clinical/demographic characteristics from 1682 Brazilian individuals referred to a private laboratory to undergo genotyping with a cancer hereditary risk multigenic panel (Table 1).

**Table 1.**
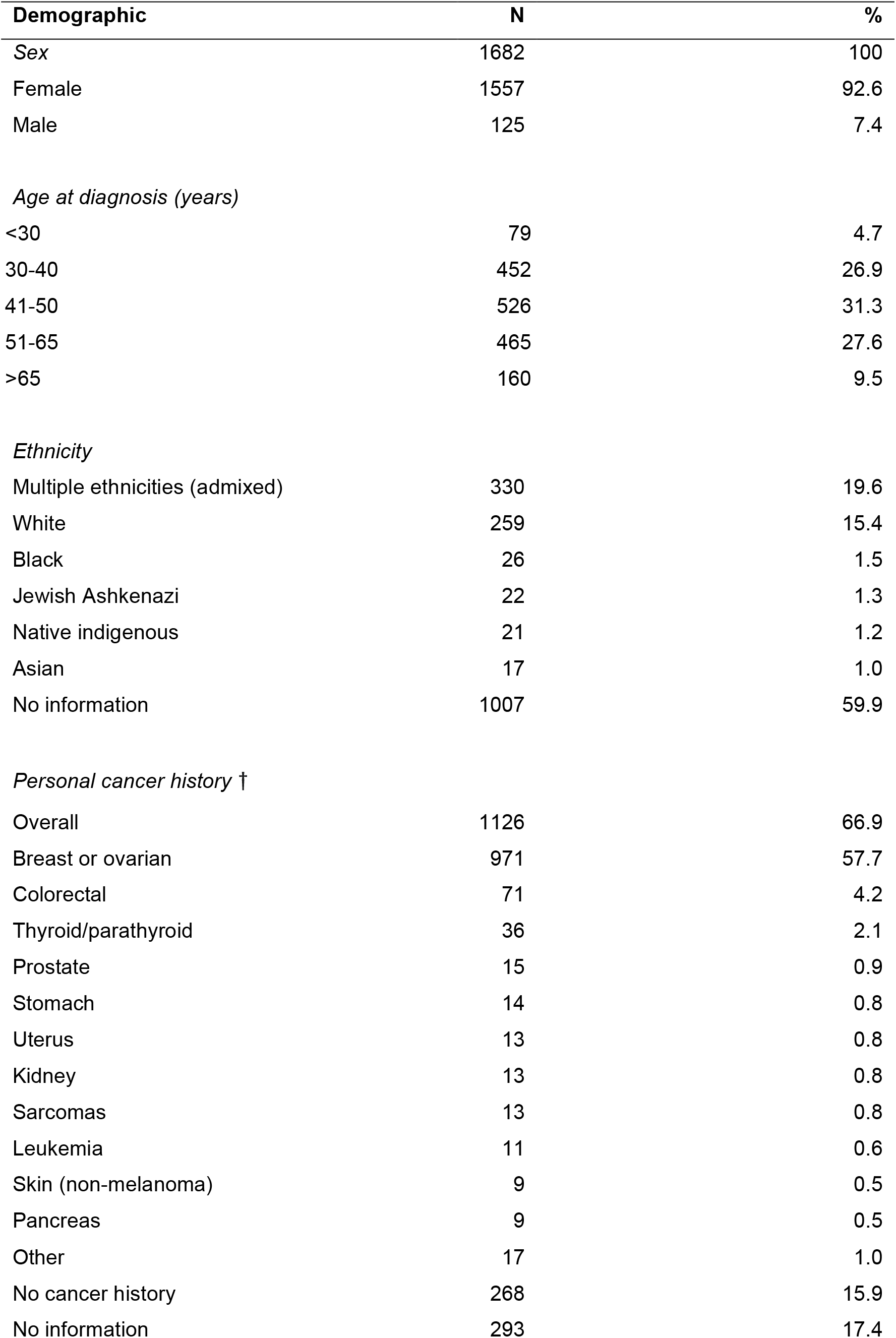
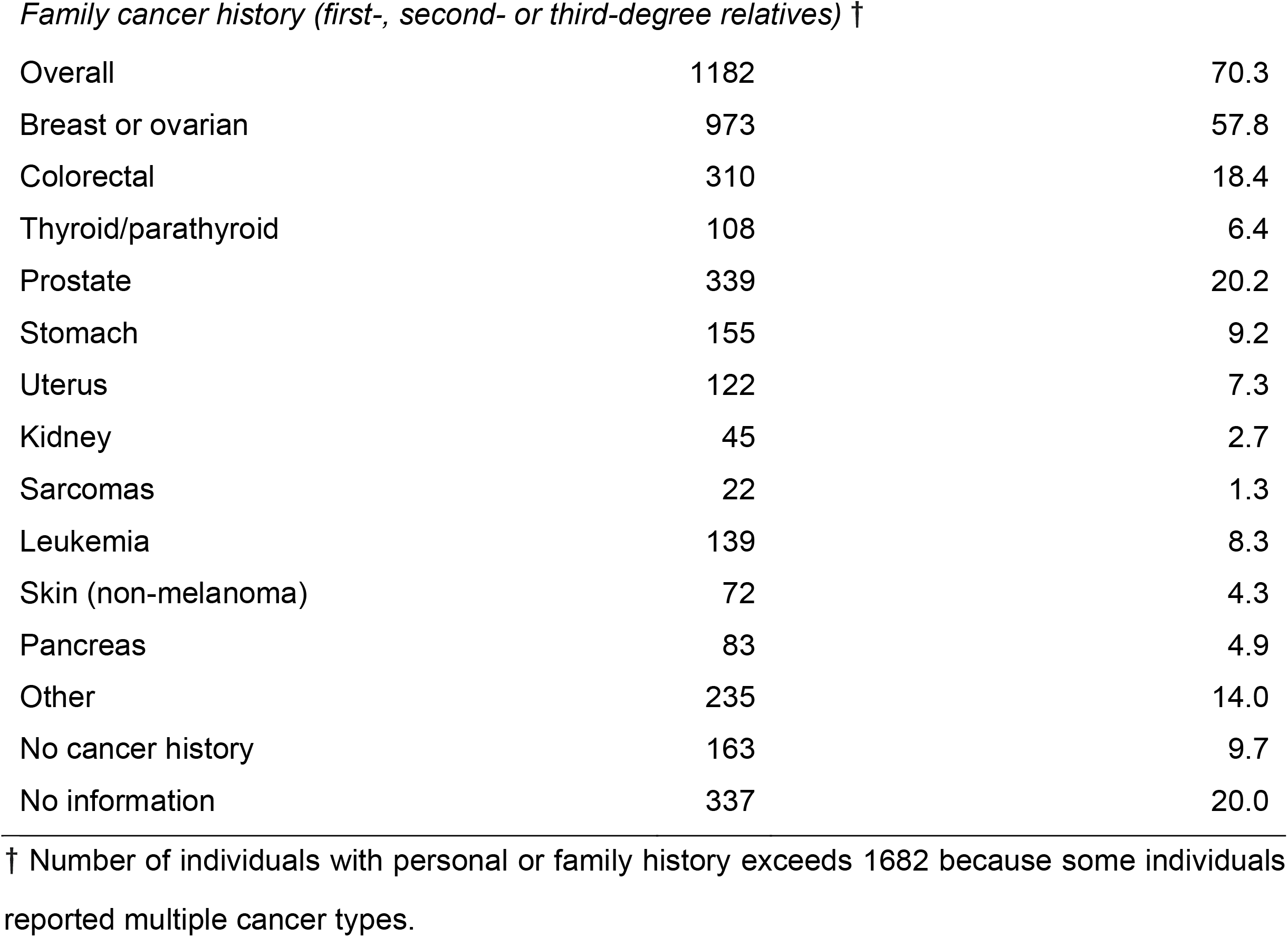
Demographic and clinical characteristics of Brazilian individuals tested genotyped with NGS hereditary cancer panel.

| Demographic | N | % |
| --- | --- | --- |
| Sex | 1682 | 100 |
| Female | 1557 | 92.6 |
| Male | 125 | 7.4 |
| *Age at diagnosis (years)* |  |  |
| <30 | 79 | 4.7 |
| 30-40 | 452 | 26.9 |
| 41-50 | 526 | 31.3 |
| 51-65 | 465 | 27.6 |
| >65 | 160 | 9.5 |
| *Ethnicity* |  |  |
| Multiple ethnicities (admixed) | 330 | 19.6 |
| White | 259 | 15.4 |
| Black | 26 | 1.5 |
| Jewish Ashkenazi | 22 | 1.3 |
| Native indigenous | 21 | 1.2 |
| Asian | 17 | 1.0 |
| No information | 1007 | 59.9 |
| *Personal cancer history †* |  |  |
| Overall | 1126 | 66.9 |
| Breast or ovarian | 971 | 57.7 |
| Colorectal | 71 | 4.2 |
| Thyroid/parathyroid | 36 | 2.1 |
| Prostate | 15 | 0.9 |
| Stomach | 14 | 0.8 |
| Uterus | 13 | 0.8 |
| Kidney | 13 | 0.8 |
| Sarcomas | 13 | 0.8 |
| Leukemia | 11 | 0.6 |
| Skin (non-melanoma) | 9 | 0.5 |
| Pancreas | 9 | 0.5 |
| Other | 17 | 1.0 |
| No cancer history | 268 | 15.9 |
| No information | 293 | 17.4 |
| *Family cancer history (first-, second- or third-degree relatives) †* |  |  |
| Overall | 1182 | 70.3 |
| Breast or ovarian | 973 | 57.8 |
| Colorectal | 310 | 18.4 |
| Thyroid/parathyroid | 108 | 6.4 |
| Prostate | 339 | 20.2 |
| Stomach | 155 | 9.2 |
| Uterus | 122 | 7.3 |
| Kidney | 45 | 2.7 |
| Sarcomas | 22 | 1.3 |
| Leukemia | 139 | 8.3 |
| Skin (non-melanoma) | 72 | 4.3 |
| Pancreas | 83 | 4.9 |
| Other | 235 | 14.0 |
| No cancer history | 163 | 9.7 |
| No information | 337 | 20.0 |

† Number of individuals with personal or family history exceeds 1682 because some individuals reported multiple cancer types.

Among the individuals, 1557 (92.6%) were women with mean age of 47.4 ± 12.3 (range 11-88) years and 125 (7.4%) were men with mean age of 51.4 ± 15.2 (range 14-91) years. Regarding ethnic ancestry, 1007 (59.9%) individuals reported unknown background, 330 (19.6%) reported having multiple ethnicities (admixed), 259 (15.4%) white, 26 (1.5%) black, 22 (1.3%) Ashkenazi Jewish, 21 (1.2%) native indigenous and 17 Asian (1.0%) (Table 1).

Personal cancer history was reported by 1126 (66.9%) individuals, with breast or ovary cancers identified in 971 individuals (57.7%); colorectal cancer in 71 (4.2%); thyroid/parathyroid in 36 (2.1%); prostate cancer in 15 (0.9%); stomach cancer in 14 (0.8%); uterus, kidney, and sarcomas in 13 each (0.8%); leukemia in 11 (0.6%); non-melanoma skin and pancreas cancers in 9 each (0.5%); and melanoma in 6 (0.4%). Other types of cancer (lung, liver, bladder, larynx etc.) were identified in 17 individuals (1.0%). Overall, 268 individuals (15.9%) did not present malignant neoplasms and for 293 individuals (17.4%) the history information was not available.

Family cancer history (considering first-, second- or third-degree relatives) was reported by 1182 (70.3%) individuals: 806 (47.9%) reported breast cancer family history; 310 (23.0%) colorectal cancers; 339 (20.2%) prostate cancer; 219 (13.0%) head and neck tumors; 172 (10.2%) lung cancer; 155 (9.2%) stomach cancer; 119 (7.1%) breast and ovarian and 48 (2.9%) ovarian cancer exclusively. No family history was reported by 163 (9.7%) individuals and 337 (20.0%) did not provide information regarding family history (Table 1, Supplementary Table 2).

### Genetic findings

Pathogenic or likely pathogenic variants were found in 305 (18.1%) of the 1682 individuals. Additionally, 1252 variants of uncertain significance (VUS) were found in 753 (44.8%) individuals (Figure 1A, Supplementary Table 3). The remaining 624 (37.1%) patients did not present any variants of clinical interest whatsoever (negatives).

**Figure 1.**
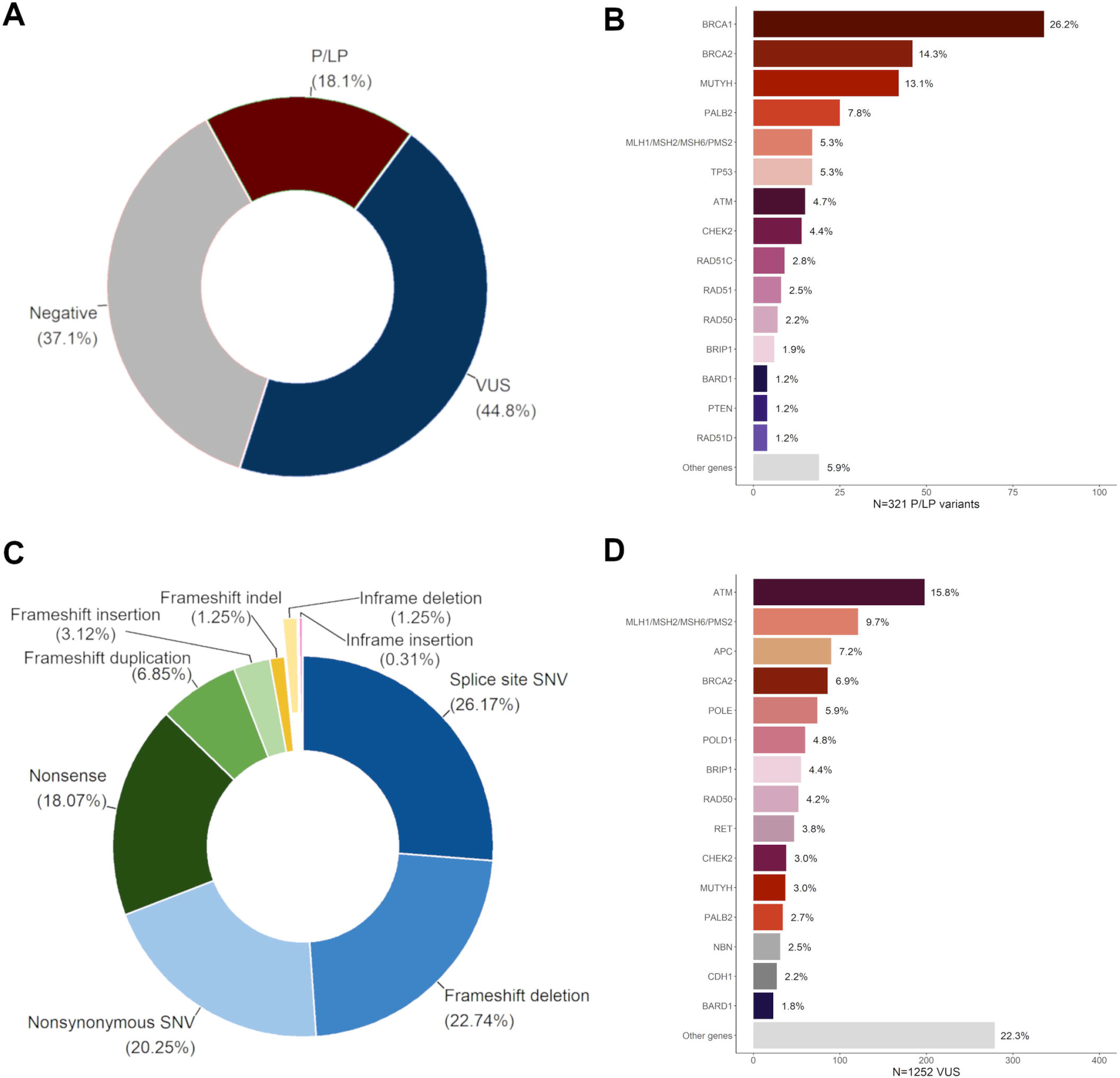
Variants profiles. (A) Distribution of patients according to genetic findings. P/LP = patients with pathogenic/likely pathogenic, VUS = patients with variants of uncertain significance (N = 1682). (B) Distribution of P/LP variants per gene. (C) P/LP variants functional annotation. (D) Distribution of VUS per gene. Lynch syndrome genes have been grouped together in (C) and (D).

The 305 individuals collectively had 321 pathogenic/likely pathogenic variant hits (corresponding to 166 unique variants) in 32 genes: *APC, ATM, BARD1, BRCA1, BRCA2, BRIP1, CDH1, CDK4, CHEK2, EPCAM, FANCC, MEN1, MITF, MLH1, MRE11A, MSH2, MSH6, MUTYH, NBN, PALB2, PMS2, POLD1, PTCH2, PTEN, RAD50, RAD51, RAD51C, RAD51D, RB1, RECQL, RET* and *TP53*.

The genes that most commonly presented pathogenic or likely pathogenic variants were *BRCA1* (84/321 = 26.2%), *BRCA2* (46, 14.3%) and *PALB2* (25, 7.8%). Lynch syndrome genes (*MLH1, MSH2, MSH6* and *PMS2*) came next with 17 (5.3%) and were tied with *TP53*, also with 17 (5.3%) hits. Next came *ATM* (15, 4.7%), *CHEK2* (14, 4.4%), *RAD51C* (9, 2.8%), *RAD51* (8, 2.5%), *RAD50* (7, 2.2%), *BRIP1* (6, 1.9%), *BARD1* (4, 1.2%), *PTEN* (4, 1.2%), *RAD51D* (4, 1.2%) and *APC* (3, 0,9%). Other genes amounted 17 (3.0%) variants (*NBN, RET*, biallelic *MUTYH, MITF, CDH1, EPCAM, FANCC, MRE11, POLD1, RB1, RECQL, MEN1, CDK4* and *PTCH2*). Moreover, monoallelic *MUTYH* variants, considered to have low penetrance, were detected in 42 (13.1%) cases (Figure 1B).

In terms of unique variants, we detected 24 in *BRCA1*, 32 in *BRCA2*, 16 in *PALB2*, 16 in the Lynch syndrome genes, eight in *TP53*, 14 in *ATM*, eight in *CHEK2*, six in *RAD51C*, nine in *RAD51*, two in *RAD50*, six in *BRIP1*, four in *PTEN*, two in *RAD51*, three in *APC*, two in *NBN*, eight in *MUTYH*, two in *CDH1* and one each in *BARD1, RET, MITF, EPCAM, FANCC, MRE11, POLD1, RB1, RECQL, MEN1, CDK4* and *PTCH2*.

Among the 321 pathogenic/likely pathogenic variant hits, 84 (26.2%) were point mutations in conserved splice sites, 73 (22.7%) were frameshift deletions, 65 (20.2%) were missense point mutations, 58 (18.1%) were nonsense point mutations, 22 (6.9%) were frameshift duplications, 10 (3.1%) were frameshift insertions, four (1.3%) were frameshift indels, four inframe deletions (1.3%), and a single inframe insertion (0.3%) (Figure 1C).

Most of the 1252 VUS hits (corresponding to 886 unique variants) were identified in *ATM* (198 hits, 15.8%), followed by *BRCA2* (86, 6.9%), *MHS6* (63, 5.0%), *BRIP1* (55, 4.4%), *RAD50* (52, 4.2%), *CHEK2* (38, 3.0%), *PALB2* (34, 2.7%), *MHS2* (31, 2.5%), *MLH1* (17, 1.4%), *TP53* (17, 1.4%), *BRCA1* (17, 1.4%). Other genes amounted to 644 (51.4%) VUS (Figure 1D).

### Individuals with multiple variants (transheterozygotes)

Among the 305 individuals with pathogenic/likely pathogenic variants, 290 (95.1%) were single heterozygotes, 14 (4.6%) presented two variants in different genes (any two of these: *APC, ATM, BRCA1, BRCA2, CDK4, CHEK2, MEN1, MUTYH, PALB2, PMS2, RAD51, RAD51C* or *TP53*) and a single patient (0.3%) presented three pathogenic variants in different genes (*BRCA1, MSH6* and *MUTYH*). The most common combination in the patients with two variants was a variant in a high-penetrance gene, such as *BRCA1* and *BRCA2*, alongside a variant with lower penetrance, such as monoallelic *MUTYH* or in *CHEK2*. No patients presented more than one pathogenic/likely pathogenic variants in the same gene (Figure 2, Supplementary Table 4). We observed a single individual *BRCA1*/*BRCA2* double heterozygote, making the double heterozygotes prevalence among *BRCA* variants carriers about 0.78% (1/129) and 0.06% (1/1682) overall. Thus, the overall transheterozygosity prevalence was about 0.89% (15/1682) in our sample.

**Figure 2.**
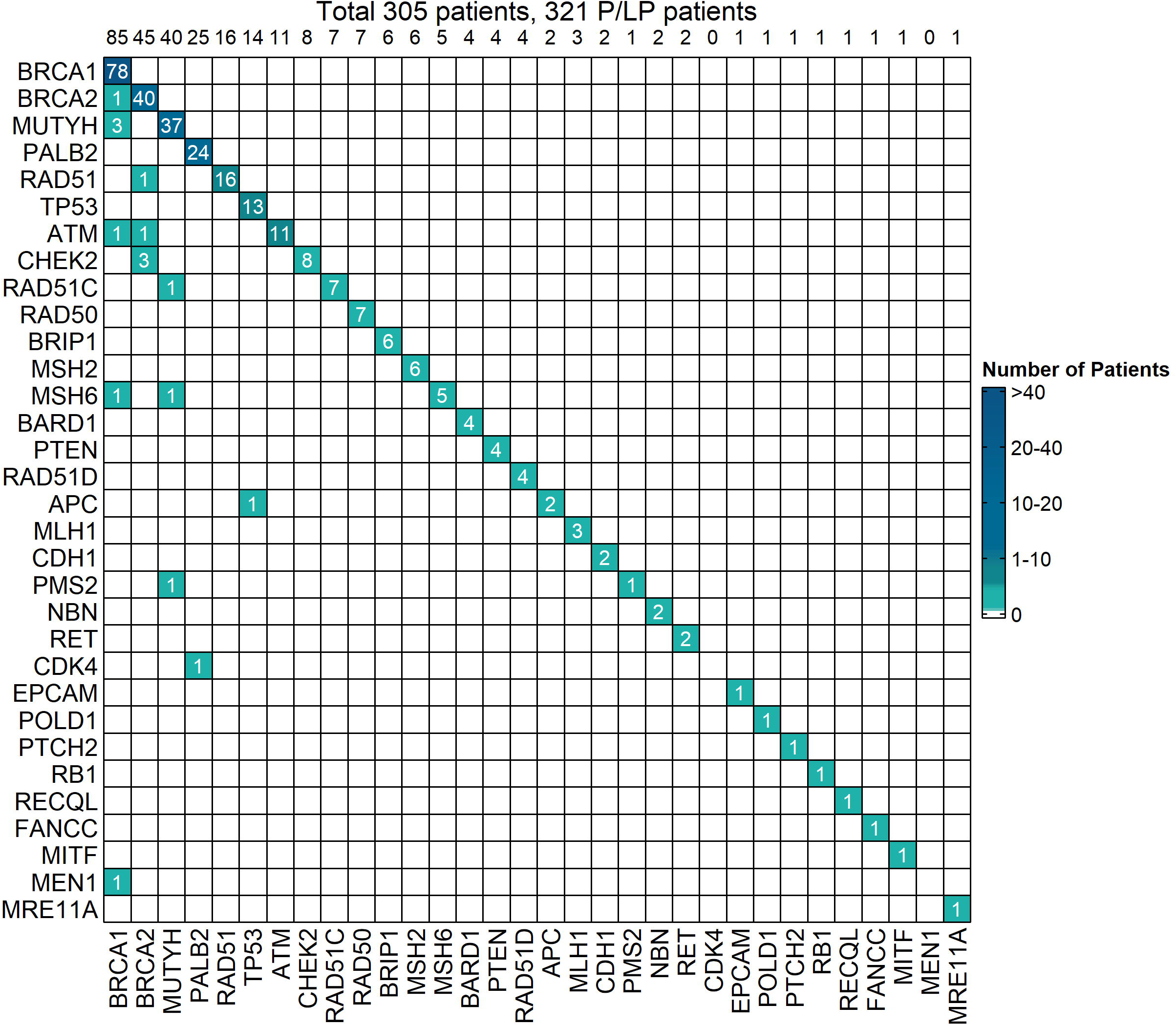
Heatmap representing the distribution of pathogenic/likely pathogenic (P/LP) transheterozygotes in the sample. The top line represents the number of patients in each column. The single heterozygotes (n=290) are distributed in the main diagonal. Transheterozygotes (n=15) are distributed in the inferior half (14 double-heterozygotes and 1 triple-heterozygote). The total number of variants are 321 (290 × 1 + 14 × 2 + 1 × 3 = 321).

### Positivity profiles according to testing criteria of the USA National Comprehensive Cancer Network and Brazilian National Health Agency

The sampled individuals were assessed for hereditary cancer testing indication according to two sets of clinical criteria: the NCCN for hereditary breast/ovarian, colorectal, stomach or pancreas cancer syndromes, version 3.2019 and 1.2020 and the criteria of the Brazilian National Health Agency (ANS). Among the 1682 individuals in study, 1008 (59.9%) met NCCN 3.2019 and 1.2020 criteria for testing, 368 (21.9%) did not meet the criteria and 306 did not have sufficient information for classification. Forty-five patients among the 305 that presented pathogenic/likely pathogenic (positive patients) were in the last group, leaving 260 classifiable patients. The true positive rate of NCCN criteria was 215/260 (82.7%) and the false negative rate was 45/260 (17.3%). The F1-measure was 33.9%. Regarding ANS criteria, 660 (39.2%) met the criteria for testing, 691 did not meet and 331 did not have sufficient information for classification. The true positive rate of ANS was 145/260 (55.7%) and the false negative rate was 115/260 (44.2%), and F1-measure was 31.5% (Table 2).

**Table 2.** NCCN 3.2019/1.2020 and Brazilian ANS testing criteria performance.

| Decision | NGS testing |  | **Row**<br>**totals** |
| --- | --- | --- | --- |
|  | Positive patients | VUS/Negative<br>patients |  |
| *NCCN 3.2019/1.2020* |  |  |  |
| At risk | 215 | 793 | **1008** |
| No risk | 45 | 323 | **368** |
| **Column totals** | **260** | **1116** | **1376** |
| Insufficient information:<br>306 |  |  |  |
| *Brazilian ANS* |  |  |  |
| At risk | 145 | 515 | **660** |
| No risk | 115 | 576 | **691** |
| **Column totals** | **260** | **1091** | **1351** |
| Insufficient information:<br>331 |  |  |  |

NCCN: sensitivity = 82.7%, specificity = 28.9%, false negative rate = 17.3%, F1-measure = 33.9%.
ANS: sensitivity = 55.8%, specificity = 52.8%, false negative rate = 44.2% F1-measure = 31.5%.

The group of 215 patients fulfilling testing criteria according to the NCCN had the following genetic profiles: 72 (36.4%) individuals having variants in *BRCA1*, 29 (14.6%) in *BRCA2*, 16 (8.1%) in *PALB2*, 14 (7.1%) in *TP53*, 11 (5.6%) in *ATM*, 9 (4.5%) in *CHEK2*, 7 (3.5%) in *RAD50* and *RAD51*, 6 (3.5%) in *RAD51C*, 4 (2.0%) in *MSH6*, 3 (1.5%) in *BRIP1, MSH2* and *RAD51D*, 2 (1.0%) in *APC, BARD1* and *CDH1* e 8 (4.0%) in other genes (*MEN1, MITF, MLH1, NBN, PMS2, POLD1, PTEN* and *RECQL*).

The group of 45 patients who did not meet the NCCN criteria had the following profiles: 9 (20.0%) individuals with variants in *MUTYH*, 7 (16.7%) in *BRCA2*, 6 (14.3%) in *BRCA1*, 5 (11.9%) in *PALB2*, 2 (4.8%) in *ATM, CHEK2, MLH1, MSH6, PTEN, RET* and *TP53*, besides 10 (23.8%) in other genes (*BARD1, BRIP1, CDK4, FANCC, MRE11A, MSH2, PMS2, PTCH2, RAD51* and *RAD51C*).

In summary, both NCCN 1.2020/3.2019 and ANS criteria failed to detect a substantial part of positive patients. The Brazilian criteria fared even worse, missing about 44% of positive patients, versus approximately 17% with NCCN 1.2020/3.2019.

### Geographic distribution of variants

We stratified our data according to each Brazilian state. The results are available in the Supplementary Table 5.

## Discussion

Cancers were the second most common cause of death worldwide in 2018, with 9.6 million deaths (1 every 6 deaths). Lung, prostate, colorectal, stomach and liver are the most common in men, whereas women are more affected by breast, colorectal, lung, cervix and thyroid ^14^. The Brazilian National Cancer Institute (INCA) estimates 450,000 new cancer cases per year (excluding non-melanoma skin cancers) for the 2020-2022 triennium ^15^.

Most tumors are sporadic, caused by somatic genetic events. However, about 10% of the patients carry germline pathogenic variants in cancer predisposition genes such as *BRCA1* or *BRCA2* ^1^. The identification of these patients is of utmost clinical importance because early diagnosis of hereditary cancer risk syndromes may improve vigilance and treatment. Therefore, genetic investigation is already a tool of modern oncology ^16^.

Next generation sequencing (NGS) technologies reduced sequencing costs allowing rapid, precise, and simultaneous analysis of a great quantity of genetic material. NGS has been used to identify genetic causes of rare diseases and cancers, in clinical and research settings. Previously, patients in investigation of HBOC or Lynch syndrome risk would be genotyped by limited, specific panels. Nowadays, as NGS is being largely utilized by clinical laboratories, the patients may undergo genotyping by multigenic panels that allow a better stratification as well as personalized oncologic management^17^.

Here, we report the results of our study, the largest conducted in Brazil, with 1682 individuals that underwent genotyping with 27- to 78-genes panels intended to detect germinative variants in cancer predisposition genes. Women are the majority in our sample (92.6%). Consequently, the most prevalent cancers were breast and ovarian (57.7%), colorectal (4.2%) and thyroid/parathyroid (2.1%).

The overall positivity rate was 18.1% (305/1682 individuals); 16.0% (269/1682) if individuals with monoallelic *MUTYH* variants are excluded. Considering just *BRCA1/BRCA2* pathogenic/likely pathogenic variants, the positivity rate was only 7.7% (129/1682 individuals). This demonstrates the importance of NGS multigene panels: it reduces the rate of false negatives, providing more information for oncologic management and prognosis.

The *BRCA* pathogenic variants positivity rate from our study seems low, but it is not unexpected since our cohort was not specifically selected for any sex, age, ethnicity, or history of cancer. Indeed, since there is significant allelic heterogeneity in *BRCA* genes, there is great variability in positivity rates in other studies ^18^. In Brazil, studies of HBOC patients observed positivity rates between 1.3% ^19^ and 27.3% ^20^. In other countries, the positivity rate among sporadic and high-risk hereditary cancer cases varied between 2.6% in the USA ^21^ and 27.9% in Japan ^22^ (reviewed in ^23^). Therefore, our positivity rate is indeed within these ranges.

We compared the mutational profile of *BRCA1* and *BRCA2* genes we observed with other 20 Brazilian studies. Among the 56 variants we detected, 33 (19 in *BRCA1* and 14 in *BRCA2*) were recurrent in other studies. The five most recurrent variants were: *BRCA1*:c.5266dupC, found by other 18 studies^18,20,24–39^, *BRCA1*:c.3331_3334delCAAG, found in other 12 studies ^18, 20, 24, 27, 29, 31, 32, 34, 35, 37, 39, 40^, *BRCA2*:c.2808_2811delACAA found in other seven studies ^18, 29–31, 34, 35, 39^, *BRCA1*: c.1687C>T found in six studies ^26, 31, 32, 34, 35^ and *BRCA1*:c.211A>G found in other five studies ^35–38, 40^.

The five most prevalent *BRCA1* mutations among the individuals sampled by our study were c.5266dupC, c.5074+2T>C, c.3331_3334delCAAG, c.1687C>T and c.211A>G. These results partially agree with a worldwide survey that included Brazilian families of women carriers of *BRCA* pathogenic variants. Rebbeck et al. (2018) ^41^ surveyed 101 Brazilian families and observed c.5266dupC, c.3331_3334delCAAG and c.1687C>T among the five most prevalent *BRCA1* mutations. Therefore, our results agree with three variants.

The five most prevalent *BRCA2* variants in our patients were c.2808_2811delACAA, c.9382C>T, c.6024dupG, c.8488-1G>A and c.5216dupA. Only c.2808_2811delACAA was in common with the Rebbeck et al. results ^41^. Interestingly, they observed c.156_157insAlu among the five most prevalent variation among 49 Brazilian families, but our results place it in sixth place, tied with other variants.

The recurrence in independent samples from Brazil indicate the variants are representative of the *BRCA1* and *BRCA2* mutational spectrum in Brazil. Indeed, the *BRCA1*:c.5266dupC and *BRCA2*:c.156_157insAlu (found in other four studies ^31, 34, 35,42^) variants are considered as founder mutations coming from European populations and may be found in the individuals from all regions of Brazil ^35^. Indeed, we detected *BRCA1:* c.5266dupC in individuals from Pernambuco and Alagoas, Northeast Brazil states, as well as in São Paulo (Southeast) and Paraná (South) and *BRCA2*:c.156_157insAlu was detected in an individual from Piauí, also a Northeast state.

We also observed some recurrent pathogenic variants in other genes: *TP53*:c.1010G>A, found in four other studies^20,38–40^, *CHEK2*:c.349A>G, found in two other studies ^38, 43^; *MUTYH*:c.1147delC ^43^; *MUTYH*:c.1187G>A ^38^, *MSH2*:c.2152C>T ^44^; *MSH6*:c.3848_3862del ^45^, *MLH1*:c.677G>A ^44^ and *PALB2*:c.2257C>T ^46^. These observations could mean that these variants are important co-players alongside the recurrent *BRCA* variants in the risk of hereditary cancers in Brazilian populations.

Transheterozygosity, i.e. heterozygosity at two different loci ^47^, is rare among patients at risk of hereditary cancers. The prevalence of double heterozygosity among *BRCA1/BRCA2* carriers ranges between 1.8% and 1.85% ^48, 49^ in the Jewish Ashkenazi populations. Among non-Ashkenazi Europeans, the prevalence is lower, between 0.22% and 0.87% ^48^. An Italian study reported 0.62% ^50^ and a Korean study reported 1.2% ^51^. Therefore, our estimate of 0.78% in the Brazilian sample seems plausible.

There has been controversy regarding the consequences over the phenotype of *BRCA1*/*BRCA2* double heterozygote. Earlier reports suggest that they do not have worse phenotypes when comparing to single-variant carriers ^52, 53^. Bell et al. (2002) ^54^ analyzed three independent samples of tumor tissue from the same *BRCA1/BRCA2* double heterozygote patient and observed that either one or other gene had suffered loss of heterozygosity (LOH) genetic events, suggesting that both genes are functionally equivalent in initiating tumorigenesis in breast tissue. Therefore, double heterozygote status would be equivalent to single-carrier status. Other case reports, case series and review of case reports also suggested that double heterozygote status was not associated with more severe phenotype ^48, 55, 56^. In contrast, Randall et al. (1998) ^57^ observed LOH in both genes in ovarian tumors, perhaps signaling that the pathogenic events in each tissue are different and not always *BRCA1* mutations are equivalent to *BRCA2* counterparts.

However, more recent large surveys have been suggesting that double heterozygote have different phenotypes when compared to single heterozygote individuals. A large survey of 8162 German families with breast/ovarian cancer cases found eight double heterozygote individuals. They observed that they were younger at the onset of their first cancer episode when compared with their single heterozygote relatives and had more severe disease ^58^. A worldwide survey of 32,295 women compared the clinical history of double heterozygote individuals with breast cancer to their single heterozygote counterparts. The same was done for ovarian cancer cases. They observed that clinical characteristics of double heterozygote individuals resemble phenotypes from *BRCA1*-only single heterozygotes, but tumor LOH microsatellite markers patterns are intermediate to the patterns of *BRCA1*-only single heterozygote and *BRCA2*-only single heterozygote. Thus, the authors suggest that the joint effect of *BRCA1*/*BRCA2* variants are compatible with an additive model ^47^. Another recent study also suggests that the double heterozygote phenotype more closely resembled that of *BRCA1* mutation carriers with poor prognosis factors and proposed a co-dominant model for the joint effects ^59^.

In summary, *BRCA1/BRCA2* double heterozygote may have early onset of disease and the phenotype is perhaps similar to the “severe end of spectrum of *BRCA1* mutation carriership” ^48^. We found a single *BRCA1/BRCA2* double heterozygote woman in our sample. She presented personal history of breast cancer. Since she is in her late 40s, her first cancer case must have happened much earlier, however, detailed data on her phenotype is currently lacking.

Literature on *BRCA*/other genes double heterozygotes is sparser. Thus, comparison with our results was difficult due to the possible different combinations of variants. For example, Sokolenko et al. (2014) ^60^ found seven digenic combinations double heterozygotes among *BRCA1* and other DNA double-strand repair genes (*BRCA1/CHEK2, BRCA1/ATM, BRCA1/BLM, CHEK2/BLM, CHEK2/ATM, NBS1/ATM, and NBS1/BLM*) in Russian patients: none were observed in our study. They did not observe differences from single heterozygote individuals. In contrast, a case series including a German *BRCA1/PALB2* double heterozygous patient had no early onset but had severe disease (multifocal triple negative ductal carcinoma) ^61^. Another case report presented a case of a double heterozygote *APC/MLH1* man with Kashmir/Egyptian ancestry, which had a history of six jejunal cancers ^62^. Therefore, double heterozygosity may have unusual effects on cancer phenotypes. However, we did not find *APC/MLH1* double heterozygous, but found an individual *APC/TP53* double heterozygote, so it is another case warranting further investigation.

Perhaps the joint effects of non-*BRCA* variants can be explained in an additive manner as well. Thus, some combinations will result in severe phenotypes while others will not, due to the specific penetrance of each variant. It seems that non-*BRCA* double heterozygotes will need to be analyzed on a case-by-case basis.

We found that NCCN 1.2020 criteria missed a substantial proportion of patients that had pathogenic/likely pathogenic variants. Other authors observed the same. We found that NCCN criteria missed a substantial proportion of patients that had pathogenic/likely pathogenic variants. Other authors observed the same. For example, Grindedal et al. (2017) ^63^ investigated *BRCA* mutations in a Norwegian breast cancer cohort and assessed some testing criteria, including NCCN, and found a false negative rate of 15.8%. Yang et al. (2018) ^64^ investigated 4196 patients genotyped with 40- to 80-genes panels and showed a false negative rate of 13.5% with NCCN criteria. Both were not much far from our estimate of 17% with NCCN 1.2020/3.2019.

Other authors proposed changes to NCCN criteria. Alemar et al. (2017) ^34^ found that adding criteria that are not included in the NCCN and ANS criteria (e.g. some ASCO criteria ^65, 66^) achieved a higher predictive value, while other authors compared other four algorithms (BOADICEA ^67, 68^, BRCAPRO, Myriad ^69^ and Manchester score ^70^) and observed that the pedigree-based BOADICEA most accurately predicted *BRCA1/BRCA2* variant carrier status in a Southeastern Brazilian population ^71^. Although NCCN criteria were imperfect, ANS criteria fared worse and a reformulation is warranted.

Recently, a panel of Brazilian experts proposed recommendations for improving testing criteria for HBOC risk patients in Brazil. Besides modifying testing criteria, the expert panel also recommended offering risk-reducing surgeries for positive patients. For negative patients, investigating both maternal and paternal lineages is warranted, so the result of models estimating cancer risk can be communicated to the patient. They also suggested that VUS should always be reported and periodically reassessed, but no urgent clinical action is justified since most VUS are constantly reclassified to benign/likely benign categories. Furthermore, patients should be contacted whenever any update in testing protocols or management options should appear ^72^.

Our study had some limitations. The NGS panel detects small deletions and duplications up to 17 base-pairs, but large deletions and duplications are not detected by this methodology. Other structural chromosomal changes, such as inversions and translocations are not detected either. If some of these changes are suspected, we recommend using methodologies like array CGH, MLPA, qPCR or FISH to confirm the variant found. Expansion variants of trinucleotide repeats, deep intronic variants or regulatory regions such as promoters are not detectable in the present test. Epigenetic changes are also not detectable by this test. At least for *BRCA* genes though, large rearrangements seem to be uncommon in Brazilian populations ^35, 42^. Moreover, further phenotype information was missing for several patients, precluding further analyses.

## Conclusion

In conclusion, we genotyped 1682 Brazilian individuals with clinical criteria of hereditary cancer syndrome from all regions of the country with NGS multigenic panels suited for the detection of germline pathogenic variants associated with cancer susceptibility, making it the largest Brazilian study of this nature to date. We observed several *BRCA1* and *BRCA2* recurrent mutations, confirming their presence in the Brazilian mutational spectrum and generated data for other 30 genes and 110 variants with modest penetrance. We also estimated the prevalence of *BRCA* as well as non-*BRCA* double heterozygotes in the Brazilian population. More studies are necessary to discover the implication of transheterozygosity over the phenotype of affected individuals.

## Data Availability

Data available within the article or its supplementary materials.

## Funding

This research did not receive any specific grant from funding agencies in the public, commercial, or not-for-profit sectors.

## Appendices

**Supplementary Table 1**. Genotyped genes.

**Supplementary Table 2**. Ancestry, genetic findings summary, testing criteria, personal and family history.

**Supplementary Table 3**. Genetic Findings.

**Supplementary Table 4**. Positivity rate per gene or combination of genes, alongside number of unique pathogenic/likely pathogenic variants detected in each gene. The variants are detailed for transheterozygotes.

**Supplementary Table 5**. Patients per Brazilian state.

## Notes

### Competing Interest Statement

The authors have declared no competing interest.

### Summary of Updates

Title and abstract updates, minor formatting changes.

